# Bullying perpetration, peer victimisation, suicidality, and their cumulative effect on preadolescents’ behaviour and brain development

**DOI:** 10.1101/2022.07.28.22278177

**Authors:** Xue Wen, Yinzhe Wang, Zaixu Cui, Xiaoqian Zhang, Runsen Chen

## Abstract

**Background:** Adolescents’ suicidality and peer bullying are rising as a serious public health concern globally. However, the neural correlates of perpetrators and the impact of different types of bullying perpetration/peer victimisation on preadolescents needs elucidation. Besides, there has been a lack of research studying the cumulative risk pattern of bullying perpetration/peer victimisation on preadolescents with suicidality.

**Methods:** We adopted a retrospective and longitudinal methodology by utilising the data from the Adolescent Brain Cognitive Development (ABCD) cohort. Firstly, participants were assigned into two groups (i.e., perpetrators vs non-perpetrators) based on their bullying perpetration experiences. Next, different types of bullying perpetration/peer victimisation (i.e., overt, relational, and reputational) were extracted to evaluate their associations with suicidal ideation (SI), non-suicidal self-injury (NSSI), and suicide attempt (SA) separately. Then, participants were further assigned into four groups (i.e., bully-uninvolved preadolescents without suicidality/NSSI history (HC group), bully-uninvolved preadolescents with SA (SA group), preadolescents with both SA and bullying perpetration (SA+P group), and preadolescents with both SA and peer victimisation (SA+V group)). We used multinomial logistic regression models, analysis of variance (ANOVA), and brain network analysis for assessing potential associations in study’s objectives.

**Findings:** In total, 9992 individuals aged 11-12 years were included in our analysis. Of these individuals, 1111 (11.1%) were classified as perpetrators, and had significantly lower bilateral cortical volume in the superior frontal sulcus (SFS), lower functional connectivity within default mode network (DMN), and lower anti-correlation between DMN and dorsal attention network (DAN) than non-perpetrators. Additionally, preadolescents’ overt, relational, and reputational perpetration/ victimisation experiences were all shown to be associated with suicidality/NSSI, despite varying strengths, while overt perpetration showed the strongest association with SA (OR 3.6 [95%CI 2.4-5.4]), followed by overt victimisation (2.4 [1.6-3.5]). Besides, the SA+V group was characterised by the highest psychopathology among four groups, while the SA+P group was characterised by the highest aggression among four groups. Lastly, lower bilateral cortical volume in the precentral gyrus (PCG) while higher cortical volume in both the lateral occipitotemporal cortex (LOTC) and posterior cingulate cortex (PCC) were found in the SA+P group when compared to SA and HC group.

**Interpretation:** Findings from the present study offered empirical evidence on the impact of bullying experience, suicidality and their cumulative risk on preadolescents’ behavior and brain development, contributing to the growing literature on discerning the impact of different types of bullying perpetration/peer victimisation on preadolescents.

## Introduction

Suicidality (i.e., suicidal ideation and suicidal attempt) and non-suicidal self-injury (NSSI) among adolescents are growing public health concerns,^1,2^ with long-lasting negative effects on children’s behavioural and neural development.^3–6^ Specifically, the aggregate lifetime prevalence was 14.2–22.7% for suicidal ideation (SI), 4.7–7.7% for suicidal attempt (SA), and 16.9–28.4% for NSSI in children and adolescents.^7^ Besides, ample research evidence has underscored that peer bullying, defined as a misuse of power in which one person (perpetrator) engages in repeated aggression against another (victim) that is intentional and involves an imbalance of power,^8^ has deleterious psychosocial impacts on children’s suicidality, psychopathology, and criminal behaviours,^9^ with the long-term effect that can persist to later adulthood.^10^ It is alarming that the incidence rate of peer bullying has not been declining as expected on a global scale in recent years, although many efforts have been made. The prevalence of peer bullying in school-aged children was 30 – 60%,^11^ and the global prevalence was 9 – 32%.^12–14^ Furthermore, one meta-regression study that aims to establish trends of peer bullying from 1998 to 2017 has not found a time trend for face-to-face peer bullying but found an increasing time trend for cyberbullying, suggesting peer bullying is still extremely relevant to children’s development as of today despite its change in format.^15^ While there has been a strong interest both in the effects of peer bullying on perpetrators and victims, the related structural and functional brain differences are not well understood,^16^ especially for perpetrators.

Determined by the intention embedded in the perpetrator’s actions, bullying perpetration/peer victimisation can be overt, relational, and reputational. Overt perpetration is frequently manifested in the perpetrator’s actions to physically damage or threat of such damage towards the victim (e.g., hitting, pushing, or threatening to beat up the victim). In contrast, relational perpetration aims to incur relational damage to the victim through harming peer relationships (e.g., social exclusion). Similarly, reputational perpetration also aims to inflict damage on the victim emotionally by damaging the victim’s reputation among peers (e.g., rumour-spreading).^17–20^ Despite the differences across different types of bullying perpetration/peer victimisation, few studies have investigated the impact of types on their clinical characteristics.^9^ Specifically, previous research has already pointed out that bullying perpetration/peer victimisation could impose significant risks for SI, NSSI, and SA,^21–25^ while whether the relationship between bullying perpetration/peer victimisation and suicidality/NSSI varies from different bullying types (i.e., overt, relational, reputational) remains unexplored.

Although the close relationship between peer bullying and suicidality has been widely established, there has been a dearth of research exploring the cumulative risk pattern of peer bullying on suicidal preadolescents. In other words, whether peer bullying itself exists as a specific risk factor in suicidal preadolescents and brings about specific negative effects on their behaviour and brain development. As implied in two longitudinal studies, compared to bully-uninvolved suicidal adolescents, bully-involved suicidal adolescents had a significantly higher risk of depression and suicide during the 2-year follow-up,^26^ and even performed more functional impairment during the 4-year follow-up.^27^ Meanwhile, compared to bully-involved non-suicidal adolescents, only bully-involved suicidal adolescents developed later depression or suicidality and reported more psychiatric problems during the 4-year follow-up.^27^ However, it remains unknown whether the cumulative risk of peer bullying on suicidal adolescents is already evident at the baseline phase and whether this effect could extend to their brain development. Clarification of this issue may suggest the need for early intervention for bully-involved suicidal preadolescents while preventing its detrimental impact on preadolescents’ development.

Therefore, there are gaps in the existing literature that can be addressed, which include elucidating the neural correlates of bullying perpetration, the differences between peer bullying and suicidality/NSSI across bullying types, and the cumulative risk pattern of peer bullying on preadolescents with suicidality. To bridge these gaps, we aimed to 1) explore the influences of bullying perpetration on preadolescents’ brain structure and function; 2) discover the relationship between overt perpetration/victimisation, relational perpetration/victimisation, reputational perpetration/victimisation, and suicidality/NSSI, respectively; 3) elaborate the cumulative risk pattern of bullying perpetration/peer victimisation on suicidal preadolescents’ behaviour and brain development.

## Methods

### Data sources

We included data from the Adolescent Brain and Cognitive Development (ABCD), “Curated Annual Released 4.0 version”. Data for all included measures were collected at the 2-year follow-up assessment for 10,414 participants between July 2018 and January 2021, with the exceptions of demographic data being collected at baseline (sex, race/ethnicity) and data of suicidality/NSSI history being collected from baseline to 2-year follow-up. The written informed consent from caregivers and verbal assent from children were obtained. All procedures were approved by the centralised institutional review board (IRB) from the University of California, San Diego, and each study site obtained approval from its local IRB.

### Measurements

The Peer Experiences Questionnaire (PEQ) was used to assess whether the child has either experienced overt, relational, or reputational victimisation from peers or perpetrated overt, relational, or reputational aggression towards peers.^28^ Each of these six domains was scored on a 5-point Likert scale ranging from 1 (“Never”) to 5 (“A few times a week”). Summary scores of bullying perpetration/peer victimisation were calculated by summing up their corresponding domain scores. Furthermore, the scores of each domain (i.e., overt perpetration/victimisation, relational perpetration/victimisation, reputational perpetration/victimisation, and overall perpetration/victimisation) were dichotomised as above the top decile score or below.^25^ The present study only used data from the 2-year follow-up from the ABCD database since relevant data were only collected in that year.

The youth-report version of the suicide module from the computerised Kiddle Schedule for Affective Disorders and Schizophrenia (KSADS, Lifetime version) was used to assess children’s past and current SI, NSSI, and SA.^29^ In order to include participants with a history of suicidality/NSSI as comprehensively as possible, three waves of data (i.e., baseline, 1-year follow-up, and 2-year follow-up) were all included to define participants’ suicidality history, while past and current diagnoses were collapsed into a single binary measurement. Participants who reported any past or current SI, NSSI, or SA from baseline to 2-year follow-up were included in our further analyses.

The parent-report version of the Child Behavior Checklist (CBCL for ages 6 to 18) was used to assess preadolescents’ psychopathology.^30^ It is a 3-point Likert scale ranging from 0 (“not true”) to 2 (“very true”). A higher total score reflects more psychopathological behaviours in preadolescents. In addition, two items related to suicidality and NSSI were selected as screening questions (i.e., “Deliberately harms self or attempts suicide” and “Talks about killing self”). Data from the 2-year follow-up were used for further analysis.

The parent version of the Early Adolescent Temperament Questionnaire-Revised (EATQ-R for ages 9 to 15) was used to assess temperament of preadolescents.^31^ It is a 5-point Likert scale ranging from 1 (“Almost always untrue”) to 5 (“Almost always true”). Herein, the aggression-hostile dimension was selected in our analysis, with higher scores reflecting more hostile and aggressive actions. Data from the 2-year follow-up were used for further analysis.

All children underwent structural Magnetic Resonance Imaging (sMRI) and resting-state functional Magnetic Resonance Imaging (rsfMRI) according to standardised protocols. The T1-weighted images acquired from 21 sites were processed at the Data Analysis, Informatics, and Resource Center (DAIRC) of the ABCD study.^32,33^ FreeSurfer v5.3 (http://surfer.nmr.mgh.harvard.edu) was used to process the locally acquired T1-weighted images and to estimate regional cortical volume. According to the Desikan-Killiany atlas, the cerebral cortex was parcellated into 74 regions in total.^34^ Data from the 2-year follow-up were analysesed. As for rsfMRI data, regions of interest were grouped together according to Gordon parcellation accompanied by subcortical and cerebellar atlases.^35^ In total, 91 cortical (78 inter- and 13 intra-network correlations) network correlations were computed. Only scans that passed the quality control for sMRI or rsfMRI were used for corresponding analyses.

Sociodemographic information, including preadolescents’ age, sex, race/ethnicity, and data acquisition site, was collected. For rsfMRI analysis, handedness and meanmotion were further considered as covariates. Detailed information related to all the used variables, including the variables used for quality control, is shown in Appendix p1.

### Statistical Analysis

All statistical analyses were performed using SPSS (version 26.0) and MATLAB (version 2022a). We use the list-wise deletion method to handle the missing data. In total, 9992 preadolescents were involved in the analyses (see Figure 1).

**Figure 1.**
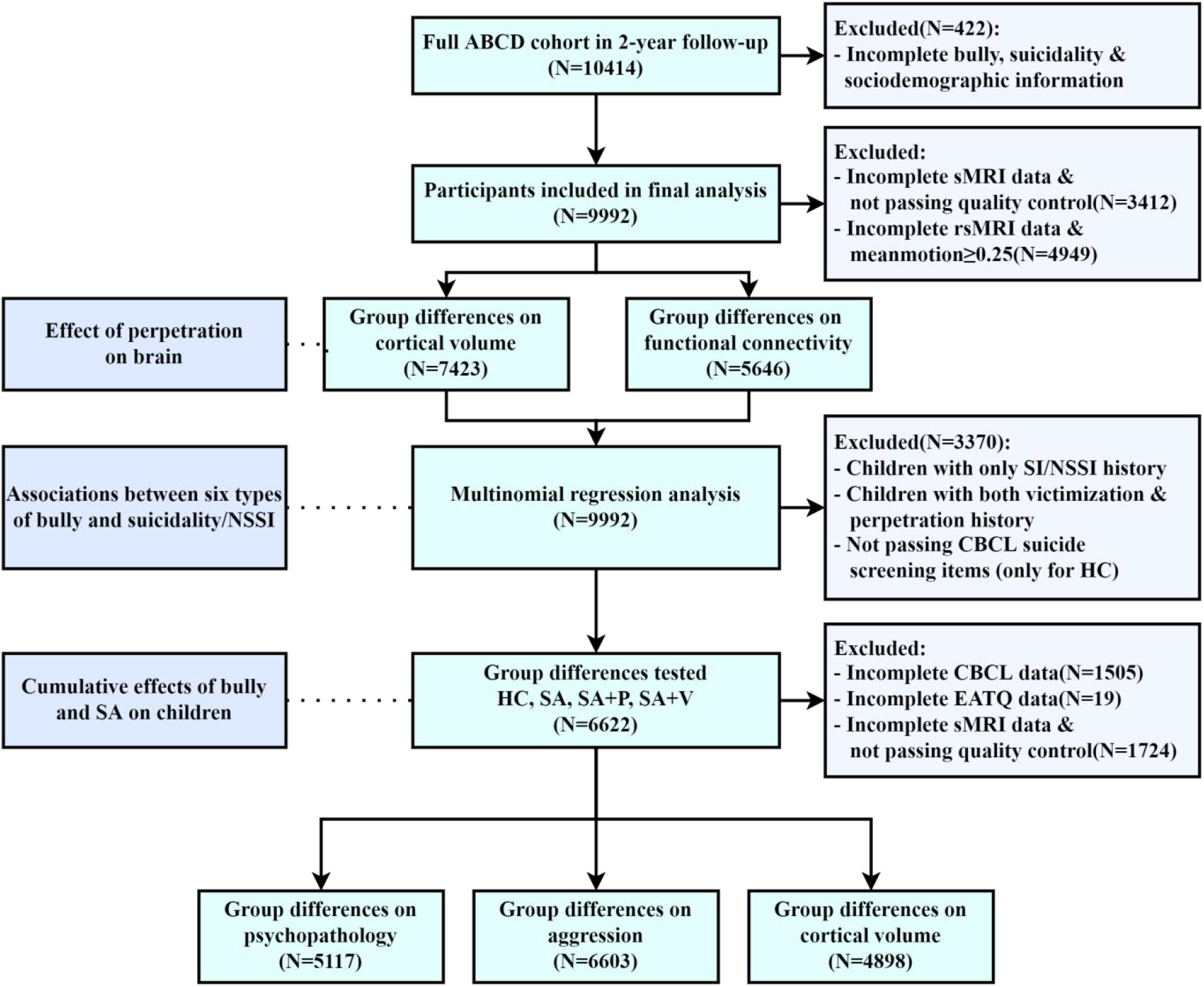
An overview of the number of participants entered into this observational analysis is shown in the workflow.

First of all, the effects of bullying perpetration on preadolescents’ brain structure and function were examined. For magnetic resonance imaging (MRI) analyses, only 7423 scans that passed the quality control were taken into the subsequent analyses. Additionally, for resting-state magnetic resonance imaging (rsMRI) analyses, 5646 qualified scans (meanmotion<0.25) were included. Controlling for the covariates above, we used multivariate analysis of variance (MANOVA) to estimate the between-group differences (Perpetrator versus Non-perpetrator) in bilateral cortical volume (across 74 regions) and large-scale resting-state functional connectivity (RSFC, across 91 connections). The false discovery rate (FDR) was further used to avoid the potential bias arising from multiple comparisons.^36^ A series of post hoc analyses were also conducted to determine the associations of the significant brain outcomes with preadolescents’ severity of perpetration.

Secondly, the associations between bullying perpetration/peer victimisation and suicidality (i.e., SI & SA)/NSSI were investigated by multinomial logistic regression models, in which the independent variables were different types of bullying perpetration/peer victimisation (i.e., overt, relational, and reputational), while the dependent variable was child-reported suicidality/NSSI.

Thirdly, four subgroups were formed to address the cumulative risk of bullying perpetration/peer victimisation on suicidal preadolescents, including bully-uninvolved preadolescents without suicidality/NSSI history (HC, N=6691), bully-uninvolved preadolescents with only SA (SA, N=106), preadolescents with both SA and bullying perpetration (SA+P, N=38), and preadolescents with both SA and peer victimisation (SA+V, N=24). In the HC group, 231 subjects were further excluded for failing the CBCL’s suicide screening questions (HC, N=6454). Then, frequencies and means (SDs) were reported for descriptive purposes. Meanwhile, χ^2^ tests or *t* tests were used for comparisons, as appropriate. Subsequently, group differences in psychopathology and aggression were tested to address the question of whether the cumulative risk of bullying perpetration/peer victimisation on suicidal preadolescents could lead to a worse effect on their clinical characteristics. The univariate analysis of covariance (ANCOVA) was used to compare the group differences over psychopathology and aggression, and then pairwise comparisons were further conducted.

Finally, the cumulative influence of bullying perpetration/peer victimisation on suicidal preadolescents’ brains was explored. For sMRI analyses, only 4898 scans that passed the quality control were taken into the final analyses. Multivariate analysis of covariance (MANCOVA) and FDR correction was used to estimate the between-group differences in cortical volume across all the 74 cortical regions.

## Results

### Effects of bullying perpetration on preadolescents’ brain structure and function

First of all, we aimed to elucidate the neural correlates of perpetration, including changes in their cortical volume and large-scale RSFC. Results after screening revealed that 1111(11.1%) preadolescents were classified as perpetrators, while the remaining 8881 preadolescents were classified as non-perpetrators. Notably, perpetrators have shown significantly lower bilateral superior frontal sulcus volume compared with non-perpetrators (*F*(1, 7417)=12.55, *p<*0.001). Meanwhile, lower RSFC (*F*(1, 5638)=14.62, *p<*0.001) within default mode network (DMN) and lower anti-correlation (*F*(1, 5638)=11.79, *p*=0.001) between DMN and dorsal attention network (DAN) was observed in perpetrators (see Figure 2). In the post hoc analyses, preadolescents’ severity of perpetration was negatively correlated with their bilateral superior frontal sulcus volume (β=-0.03, *p*=0.008) and within-DMN RSFC (β=-0.04, *p*=0.003), while positively correlated with their DMN-DAN RSFC (β=0.03, *p*=0.021).

**Figure 2.**
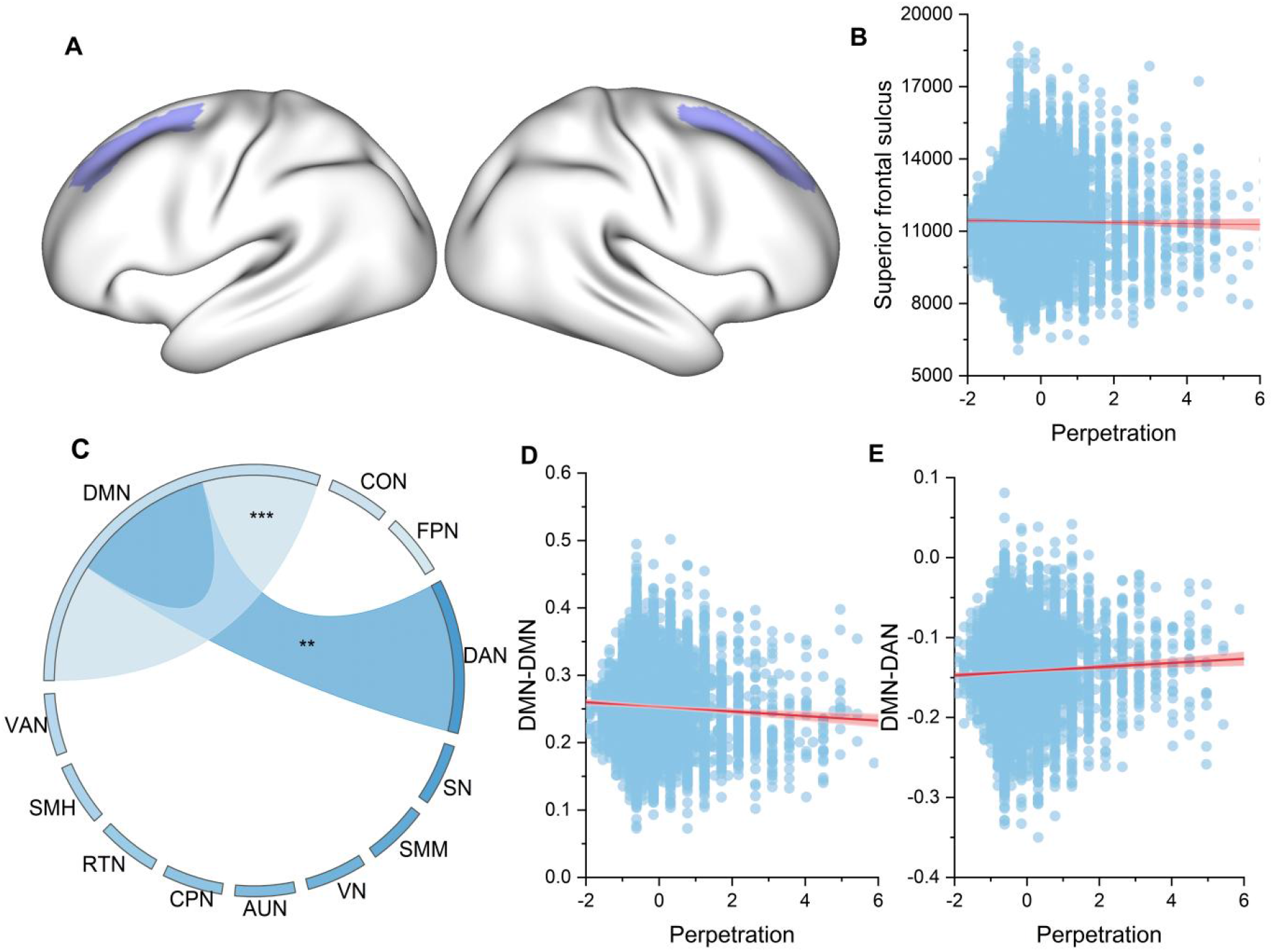
Effects of perpetration on children’s cortical volume and DMN-related functional connectivity. DMN=default mode network; CON=cingulo-opercular network; FPN=fronto-parietal network; DAN=dorsal attention network; SN=salience network; SMM=sensorimotor mouth network; VN=visual network; AUN=auditory network; CPN=cingulo-parietal network; RTN=retrosplenial temporal network; SMH=sensorimotor hand network; VAN=ventral attention network.

### Association of different types of bullying perpetration/peer victimisation with suicidality/NSSI

Next, we sought to delineate the association between six different types of bullying perpetration/peer victimisation and suicidality/NSSI (see Table 1). Controlling for demographics, we found that all six types of bullying perpetration/peer victimisation were associated with SI in smaller effect sizes (OR [1.3-1.9]), and associated with NSSI in similar effect sizes (OR [1.5-1.8]) except for relational victimisation. In contrast to SI and NSSI, overt perpetration (OR 3.6 [95% CI 2.4-5.4]; P <0.001), relational perpetration (1.5 [1.0-2.2]; P =0.033), overt victimization (2.4 [1.6-3.5]; P < .001), relational victimization (1.9 [1.2-2.8]; P =0.003), and reputational victimization (2.0 [1.4-3.0]; P <0.001), were associated with SA in much higher effect sizes, whereas reputational perpetration was not (1.3 [0.9-2.0]; P = 0.187).

**Table 1.**
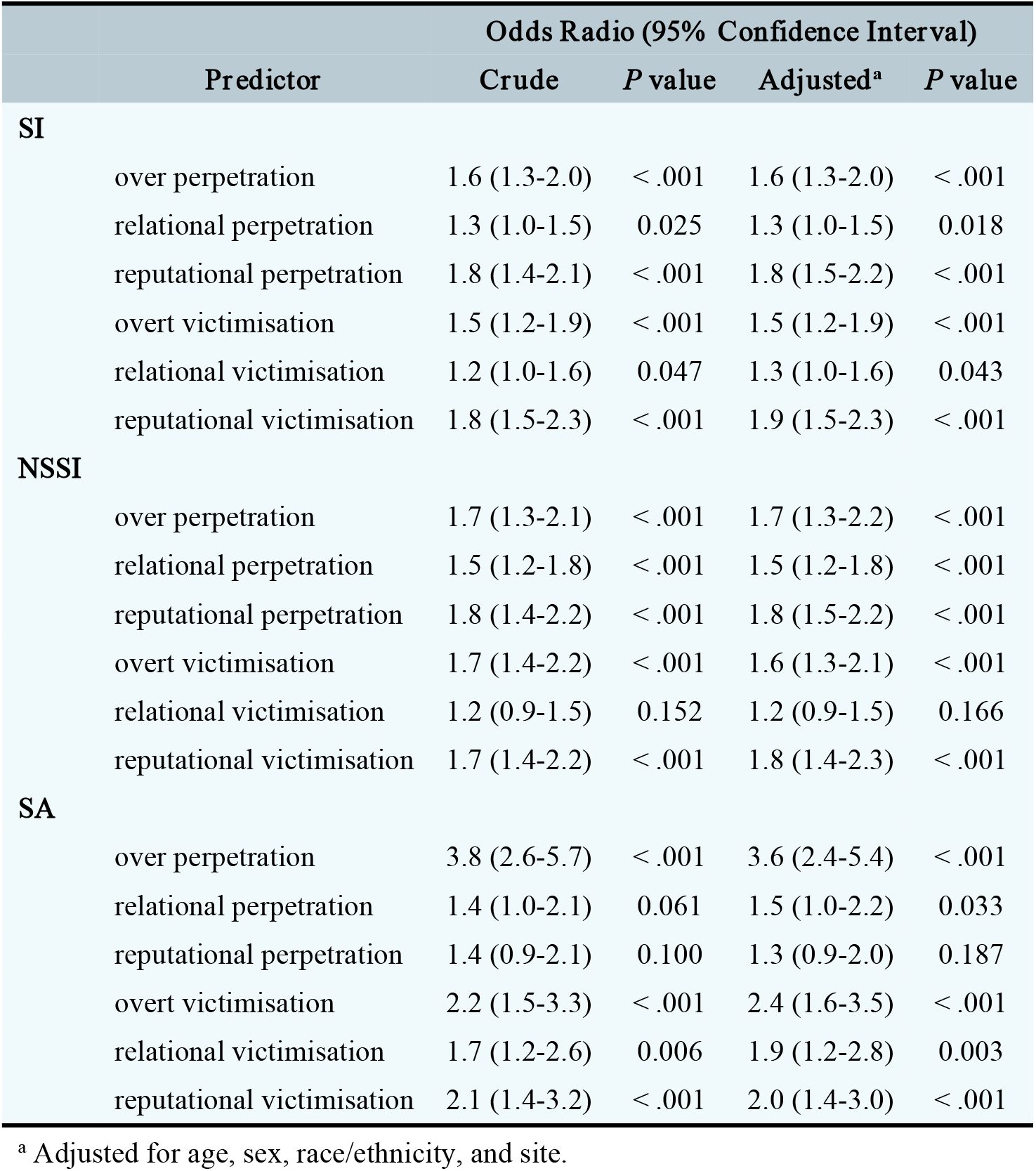
The associations of overt perpetration/victimisation, relational perpetration/victimisation, and reputational perpetration/victimisation with suicidality/NSSI

### The cumulative risk pattern of bullying perpetration/peer victimisation on suicidal preadolescents’ clinical characteristics

The demographic details of four subgroups are summarised in Table 2. Significant demographic differences in race/ethnicity (*F*(3, 6618)=5.58, p=0.001), psychopathology (*F*(3, 5109)=55.20, p<0.001) and aggression (*F*(3, 6595)=26.79, p<0.001) were found among four subgroups (HC, SA, SA+P, and SA+V). Notably, preadolescents with both SA and perpetration (Mean=58.0, SD=2.0) and preadolescents with both SA and victimisation (Mean=59.1, SD=2.4) achieved significantly higher scores in psychopathology than preadolescents with only SA (Mean=51.7, SD=1.2). Additionally, preadolescents with both SA and bullying perpetration (Mean=51.7, SD=1.2) achieved significantly higher scores in aggression than preadolescents with only SA (Mean=51.7, SD=1.2), as shown in Figure 3A and Figure 3B.

**Figure 3.**
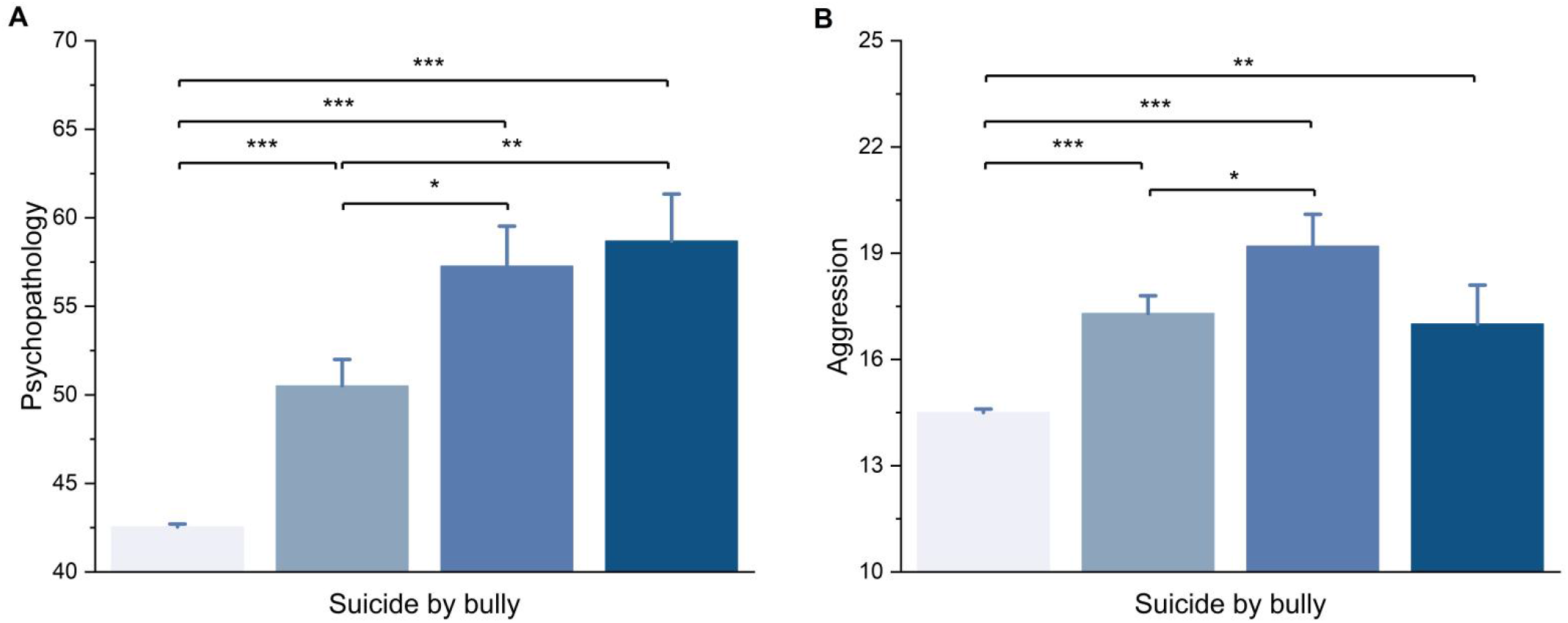
The cumulative influences of SA and bullying perpetration/peer victimisation on children’s psychopathology and aggression.

**Table 2.**
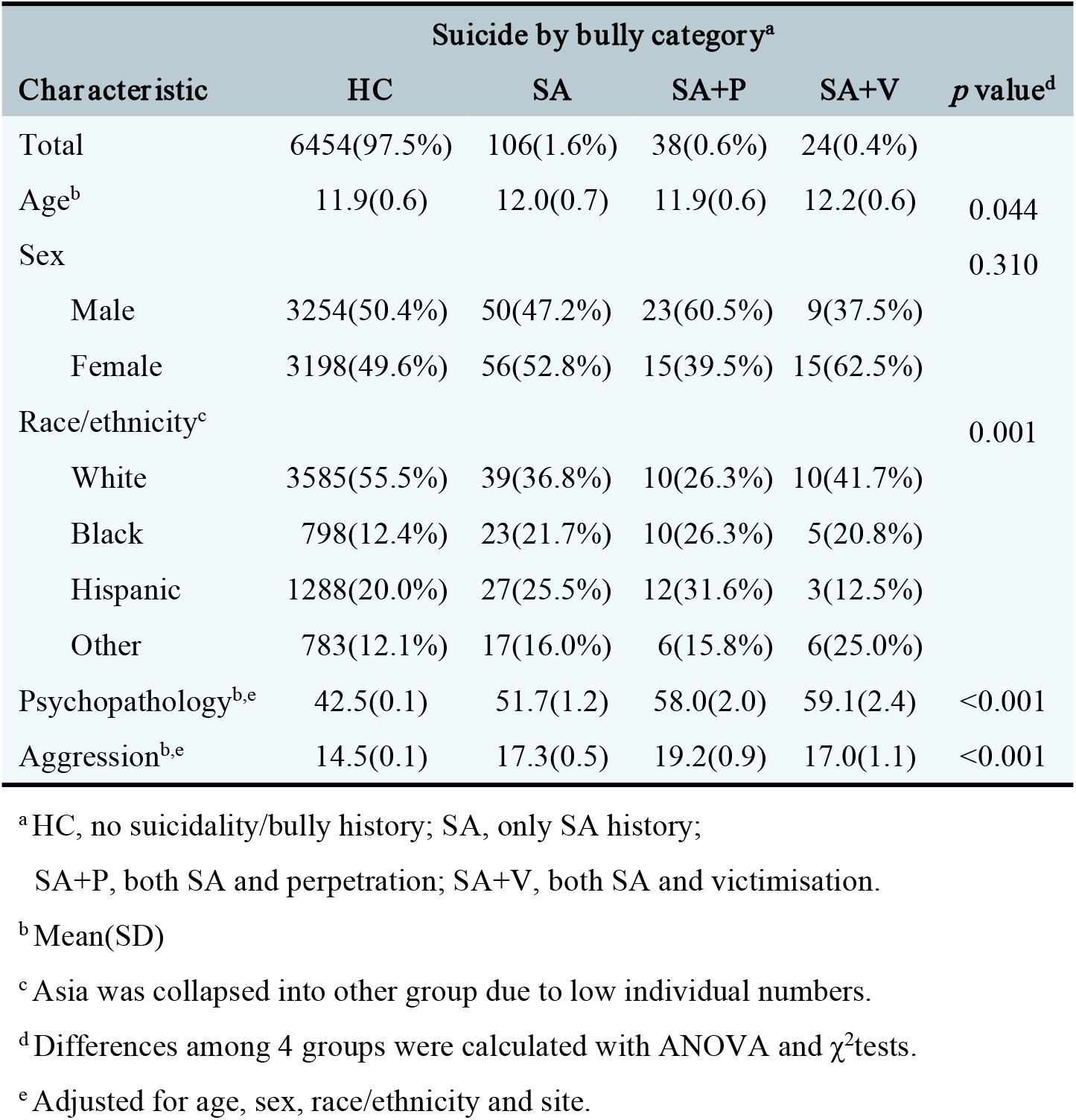
Sociodemographic and behavioral characteristics of the study participants

### The cumulative risk pattern of bullying perpetration/peer victimisation on suicidal preadolescents’ brain structures

As shown in Figure 4, significant differences in five regions’ cortical volumes (out of 74 regions) across the four subgroups were found, including paracentral lobule and sulcus (*F*(3, 4890) = 5.45, P_FDR_ = 0.019), middle-posterior part of the cingulate gyrus and sulcus (*F*(3, 4890) = 5.20, P_FDR_ = 0.019), posterior-dorsal part of the cingulate gyrus (*F*(3, 4890) = 5.18, P_FDR_ = 0.019), precentral gyrus (*F*(3, 4890) = 7.88, P_FDR_ < 0.001), and lateral occipito-temporal sulcus (*F*(3, 4890) = 4.69, P_FDR_ = 0.044). Compared to preadolescents with only SA, preadolescents with both SA and perpetration had a significantly lower bilateral precentral gyrus volume while having a significantly higher bilateral cortical volume in the lateral occipito-temporal sulcus and middle-posterior part of the cingulate gyrus and sulcus.

**Figure 4.**
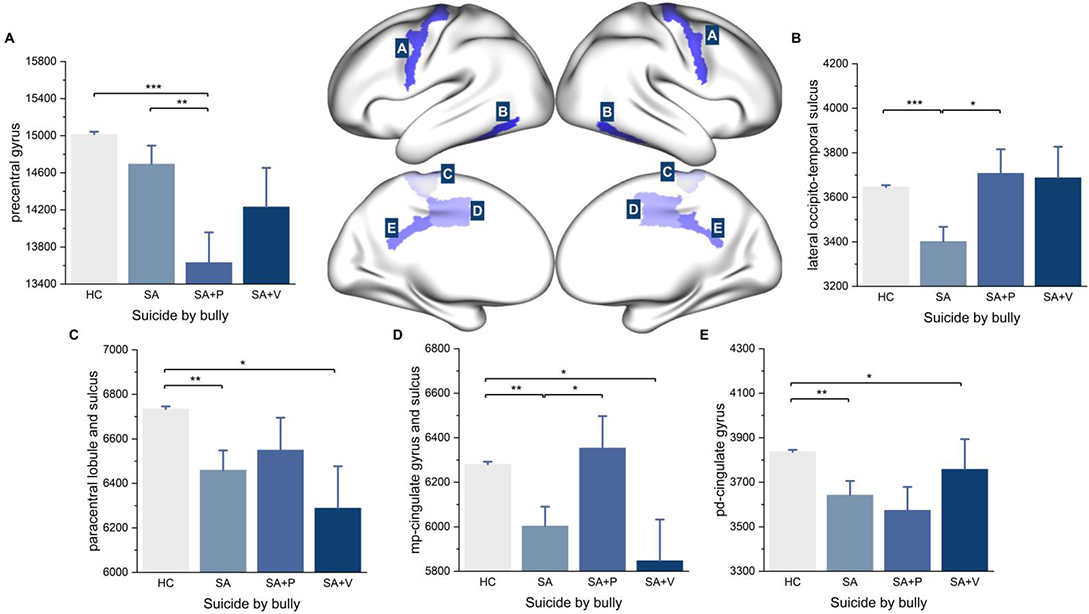
The cumulative influences of SA and bullying perpetration/peer victimisation on children’s cortical volume.

## Discussion

The current study investigated the influences of bullying perpetration on preadolescents’ brain structure and function, the differences between peer bullying and suicidality/NSSI across bullying types, and the cumulative risk pattern of bullying perpetration/peer victimisation on suicidal preadolescents. Our findings provided four clinically informative insights on 1) significantly lower superior frontal sulcus (SFS) volume, lower within-DMN RSFC, while higher DMN-DAN RSFC in preadolescent perpetrators; 2) different associations between six types of bullying perpetration/peer victimisation (i.e., overt, relational, reputational) and suicidality/NSSI in preadolescents; 3) highest psychopathology in suicidal preadolescents with victimisation, while highest aggression in suicidal preadolescents with perpetration than bullying-uninvolved suicidal adolescents than health control (i.e., bully-uninvolved non-suicidal preadolescents); 4) lower precentral gyrus (PCG) volume, while higher cortical volume in both the lateral occipitotemporal cortex (LOTC) and posterior cingulate cortex (PCC) in suicidal preadolescents with perpetration when compared to bullying-uninvolved suicidal adolescents and health control.

The first finding of the current study has filled the gap by highlighting altered brain structure and RSFC in preadolescent perpetrators. Indeed, perpetrators performed impairment in decision-making and were always accompanied by the tendency for “myopia for the future”.^37^ Specifically, perpetrators preferred unfavorable behaviours with immediate gains (e.g., venting emotions, a sense of power, being popular) to punishment and losses in the long run (e.g., ethical pressure, external criticisms). Meanwhile, extensive evidence suggests that the dorsal-lateral prefrontal cortex (dlPFC) was the seat of perceptual decision-making.^38–41^ As a portion of dlPFC, the SFS was also closely linked to this advanced cognitive process.^42–44^ Additionally, a transcranial magnetic stimulation (TMS) study targeted at human left SFS also validated the causal relation between SFS and decision-making.^44^ Thus, the decreased SFS volume reported in the current study might provide novel neurobiology evidence for the clinical intervention for preadolescent perpetrators.

Furthermore, DMN has played a crucial role in internally directed cognition, especially for future imaging events.^45^ The lower within-DMN RSFC found in our study might explain perpetrator’s ignoration to the long-term negative impact of their aggression behaviours. Notably, the DMN-DAN anticorrelation presented optimal allocation of the cognitive resource by the brain, thus was always considered a helpful property in the human brain.^46^ However, decreased DMN-DAN anticorrelation was linked to multiple neurological and psychiatric disorders (e.g., major depression disorder, conduct disorder, attention-deficit hyperactivity disorder, schizophrenia).^47–51^ Combined with the decreased DMN-DAN anticorrelation in perpetrators discovered in our study, it might partly explain their significantly increased vulnerability to developing future psychiatric disorders. ^52^ The results in the current study provided a novel neurobiological basis for preadolescent perpetrators underlying their behavioural mechanisms.

The current study found different associations with suicidality/NSSI across all investigated types of bullying perpetration/peer victimisation. Indeed, despite the fact that different parties involved during bullying (i.e., perpetrator and victims) would all face the risk of mental health disorders in their later life, their risk profiles could be different.^9^ As further accentuated in that study’s findings, although belonging to the same role in bullying, the different types (i.e., overt, relational, reputational) would have different effects on individuals, suggesting the moderating effect of bullying perpetration/peer victimisation types. Contrary to our research, previous studies implicated that physical victimisation was only associated with SI, relational victimisation was only associated with SA,^53^ and aggressive problems brought by relational victimisation could be more serious than physical victimization,^54^ while a stronger association between overt victimisation and suicidality/NSSI than relational victimisation was found in our study. Besides, the strongest association between overt perpetration and SA was also found, suggesting a high-risk subgroup in bully-involved preadolescents. Nonetheless, due to the scarcity of research in this field,^9^ future studies should focus more on the behavioural mechanisms underlying different types of bullying perpetration/peer victimisation to elucidate their different clinical characteristics.

The third major finding is that we found the highest psychopathology in suicidal preadolescents with victimisation while the highest aggression among suicidal preadolescents with perpetration. Our finding on psychopathology among bullying-involved suicidal preadolescents aligns with previous research stating that mental health conditions (e.g., depressive mood) play a mediating role in the association between bullying and suicidality.^24^ Furthermore, compared to victims, perpetrators would face a higher risk of externalising disorders (e.g., oppositional defiant disorder) and criminality (e.g., assault and illicit drug misuse) due to their aggressive, oppositional, and delinquent behavioural patterns formed during the early development stage.^9,55^ On the contrary, compared to perpetrators, victims would face a higher risk of internalising disorders (e.g., generalised anxiety disorder and major depressive disorder).^9^ Combined with this findings, we consider it would be reasonable to speculate that bullying-involved suicidal preadolescents would face an even more difficult developmental stage transition (i.e., from preadolescence to adolescence and adulthood), resulting in their worsening psychopathological state. Regarding our finding on the highest aggression among suicidal preadolescents with perpetration, numerous studies have suggested that bullying perpetration was a constant, repeated, and intentional violent and aggressive act against a victim.^56,57^ The aggression among suicidal preadolescents with perpetration could be explained by their intention to exploit the power differential between them and their victims. Another possible explanation behind their aggression could be that their low self-esteem serves as a mediating role between the association of aggressive bullying perpetration and suicidality. However, despite the plausible explanation, results from one meta-analysis examining the association between self-esteem and bullying perpetration/peer victimisation have shown that the association between perpetrator’s self-esteem and aggressive perpetration was weak and postulated that other etiological factors could be involved and needed to be found by future studies.^58^ Nevertheless, since past studies have shown that self-esteem could be influential among suicidal preadolescents,^59,60^ the interplay between self-esteem, suicidality, and aggressive bullying perpetration should be further examined by future studies.

Our further work extended the clinical characteristics of suicidal preadolescents with perpetration by elucidating their altered brain structure than bully-uninvolved suicidal preadolescents and health control. Due to the association between aggression and thinner cortical thickness of the PCG in school-aged children reported in one previous study,^61^ the lowest PCG volume in suicidal preadolescents with perpetration found in our study might be explained by the highest aggression level performed in that group. A study of cognitive-behavioural therapy (CBT) on preadolescent perpetrators also implied that the fractional amplitude of low-frequency fluctuations (fALFF) of PCG is highly associated with intervention efficacy.^62^ Additionally, consistent with previous studies, individuals with SA history had significantly decreased cortical volume in PCC and LOTC.^4,63,64^ Notably, PCC might be the neural basis of an individual’s aggressive behaviours.^65^ According to the social learning theory, children often learn aggressive behaviours through observational learning and imitate aggressive role models in multiple ways (e.g., TV, video games).^66,67^ Observational learning, the key element in the social learning theory, is one of the key functions of LOTC.^68^ Meanwhile, viewing TV violence recruited children’s LOTC, and extensive viewing can cause plenty of aggressive scripts occupying the long-term memory stored in the PCC, thus contributing to the externalisation of aggressive behaviours.^69^ A fMRI study also found activation of PCC in individuals with SA history when viewing knives versus natural landscapes.^70^ The evidence above might help to explain the higher volume of PCC and LOTC in suicidal preadolescents with perpetration than bully-uninvolved preadolescents due to their frequent activations for PCC and LOTC.

Overall, one possible explanation behind the association between bullying perpetration/peer victimisation and suicidality/NSSI is the moderating effect of the perceived emotional support and family environment of the bullying perpetrators/victims.^24,71,72^ Specifically, regarding perceived emotional support, one study in China found that a higher perceived emotional support is associated with decreased perpetration/victimisation.^72^ Since the sample of the present study consists of school-age preadolescents, one of the most key emotional support sources could be teachers at school. The same study has shown that a better-perceived relationship with teachers could also lead to decreased perpetration/victimisation. Regarding the family environment, one study has found that elements of family environment, including cognitive stimulation for deficits (e.g., help from parents to solve language problems, help from parents to resolve imperfect causal understanding, parental endorsement on academic matters, and parental guidance on poor inhibitory control), emotional support and television exposure, could be influential factors in protecting grade-school age children from risks of subsequent bullying behaviours.^71^ More specifically, their findings suggest that the increase in cognitive stimulation for deficits and emotional support could decrease the risk of children’s subsequent bullying behaviour, whereas the increase in exposure to television could result in a significant risk of children’s subsequent bullying behaviour.^71^ Based on these findings, future interventions targeting bullying prevention should utilise an ecosystemic approach for bullying prevention and focus on 1) training school teachers on how to establish more stable and reliable relationships with their students, and 2) training parents in how to fill a sense of security into their children’s emotional reservoir through pragmatic measures such as psychoeducation.

A few methodologic limitations should be taken into consideration seriously. First, only self-reported information about bullying perpetration/peer victimisation was applied, while future studies would better consider peer nomination or parent/teacher reports. Second, the number of suicidal preadolescents with perpetration/victimisation was relatively low compared with bully-uninvolved suicidal preadolescents, which may lead to reduced power for our subgroup analyses. Third, only SA was considered in the subgroup analyses, while the cumulative risk pattern of bullying perpetration/peer victimisation on preadolescents with NSSI remained unexplored.

Overall, bullying perpetration had a pervasive effect on preadolescents’ brain structure and DMN-related functional connectivity. Meanwhile, overt perpetration/victimisation, relational perpetration/victimisation, and reputational perpetration/victimisation all presented varying strengths of association with suicidality/NSSI. Besides, the cumulative risk pattern of bullying perpetration/peer victimisation on suicidal preadolescents was observed both in preadolescents’ clinical characteristics (i.e., psychopathology and aggression) and brain development.

## Data Availability

Data used in the preparation of this article were obtained from the ABCD Study (https://abcdstudy.org) and are held in the NIMH Data Archive (NDA).

## Contributors

CRS, WX, ZXQ were responsible for the conception, organisation, and execution of the study. WX were responsible for the statistical analyses. WX and WYZ were responsible for the manuscript preparation. CRS, CZX, ZXQ were responsible for the manuscript revision. CRS and ZXQ and CZX were responsible for project supervision. All authors had full access to all the data in the study and confirmed their responsibility for the decision to submit it for publication.

## Acknowledgements

We thank the Adolescent Brain Cognitive Development (ABCD) participants and their families for their time and dedication to this project. Data used in the preparation of this article were obtained from the ABCD Study (https://abcdstudy.org) and are held in the NIMH Data Archive (NDA). This is a multisite, longitudinal study designed to recruit more than 10,000 children aged 9-10 and follow them over 10 years into early adulthood. The ABCD Study is supported by the National Institutes of Health (NIH)and additional federal partners under award numbers U01DA0401048, U01DA050989, U01DA051016, U01DA041022, U01DA051018, U01DA051037, U01DA050987, U01DA041174, U01DA041106, U01DA041117, U01DA041028, U01DA041134, U01DA050988, U01DA051039, U01DA041156, U01DA041025, U01DA041120, U01DA051038, U01DA041148, U01DA041093, U01DA041089, U24DA041123, U24DA041147. A full list of supporters is available at https://abcdstudy.org/federal-partners.html. A listing of participating sites and a complete listing of the study investigators can be found at https://abcdstudy.org/principal-investigators/. ABCD consortium investigators designed and implemented the study and/or provided data but did not necessarily participate in the analyses or writing of this report. This manuscript reflects the views of the authors and may not reflect the opinions or views of the NIH or ABCD consortium investigators. The ABCD repository grows and changes over time. The ABCD data used in this report came from http://dx.doi.org/10.15154/1523041. DOIs can be found at nda.nih.gov.

