## Appendix for "Bullying perpetration, peer victimisation, suicidality, and their cumulative effect on preadolescents’ behaviour and brain development"

**Supplementary Table 1. ABCD data release 4.0 variables used in current analysis**

| Scales in dataset | Variable labels in dataset | Variable labels in current report |
|---|---|---|
| pdem02 | demo_sex_v2 | Sex at birth |
| pdem02 | interview_age | Age |
| acspsw03 | race_ethnicity | Race/Ethnicity |
| abcd_lt01 | site_id_l | Site |
| abcd_peq01 | . | Bullying perpetration/peer Victimisation |
| abcd_ksad501 | . | SI, NSSI, SA |
| abcd_cbcls01 | . | Psychopathology |
| abcd_eatqp01 | . | Aggression |
| abcd_mrisdp10201 | . | Brain structure (sMRI) |
| mriqcrp10301 | iqc_t1_ok_ser > 0 | Quality control (sMRI) |
| abcd_fsurfqc01 | fsqc_qc ~= 0 | Quality control (sMRI) |
| abcd_betnet02 | . | Functional connectivity (RSFC) |
| abcd_betnet02 | rsfmri_c_ngd_meanmotion<0.25 | Quality control (RSFC) |

"." means multiple variables were used in that scale.
